# Persistent malaria transmission from asymptomatic children despite highly effective malaria control in eastern Uganda

**DOI:** 10.1101/2021.05.04.21255999

**Authors:** Chiara Andolina, John Rek, Jessica Briggs, Joseph Okoth, Alex Musiime, Jordache Ramjith, Noam Teyssier, Melissa Conrad, Joaniter I. Nankabirwa, Kjerstin Lanke, Isabel Rodriguez-Barraquer, Lisette Meerstein-Kessel, Emmanuel Arinaitwe, Peter Olwoch, Philip J. Rosenthal, Moses R. Kamya, Grant Dorsey, Bryan Greenhouse, Chris Drakeley, Sarah G. Staedke, Teun Bousema

**Affiliations:** Department of Medical Microbiology, Radboud University Nijmegen Medical Centre, Nijmegen, the Netherlands; Infectious Diseases Research Collaboration, Kampala, Uganda; Department of Medicine, San Francisco General Hospital, University of California, San Francisco, USA; Department for Health Evidence, Radboud University Medical Center, Nijmegen, the Netherlands; Department of Medicine, Makerere University College of Health Sciences, Kampala, Uganda; Department of Infection Biology, London School of Hygiene and Tropical Medicine, London, UK; Department of Clinical Research, London School of Hygiene and Tropical Medicine, London, UK

**Keywords:** malaria, transmission, falciparum, longitudinal study, gametocyte

## Abstract

**Background:** Persistent asymptomatic *Plasmodium falciparum* infections are common in malaria-endemic settings, but their contribution to transmission is poorly understood.

**Methods:** A cohort of children and adults from Tororo, Uganda was closely followed for 24 months by continuous passive surveillance and routine assessments. *P. falciparum* parasite density, gametocyte density and genetic composition were determined molecularly; mosquito membrane feeding assays were performed on samples from participants with symptomatic and asymptomatic infections.

**Findings:** From October 2017 to October 2019, we followed all 531 residents from 80 households. Parasite prevalence was 5·8% by microscopy and 17·3% by PCR at enrolment and declined thereafter. We conducted 538 mosquito feeding experiments on samples from 107 individuals. Mosquito infection rates were strongly associated with gametocyte densities of participants. Considering both transmissibility of infections and their relative frequency, the estimated human infectious reservoir was primarily asymptomatic microscopy-detected infections (83·8%), followed by asymptomatic submicroscopic (15·6%) and symptomatic (0·6%) infections. Over half of the infectious reservoir was children aged 5-15 years (56·8%); individuals <5 years (27·5%) and ≥16 years (15·7%) contributed less. Four children were responsible for 62·6% (279/446) of infected mosquitos and were infectious at multiple timepoints.

**Interpretation:** Individuals with asymptomatic infections were important drivers of malaria transmission. School-aged children were responsible for over half of all mosquito infections, with a small minority of asymptomatic children highly infectious. Demographically targeted interventions, aimed at school-aged children, could further reduce transmission in areas under effective vector control.

**Funding:** National institute of Health, Bill & Melinda Gates Foundation, European Research Council.

## INTRODUCTION

Despite global efforts to control and eliminate malaria, progress has plateaued in recent years, and in some high transmission settings malaria burden is increasing.^1^ Optimizing vector control and treatment of symptomatic cases are central to malaria control and elimination efforts. However, symptomatic malaria cases reflect only a small proportion of all *Plasmodium* infections, and many infected individuals are asymptomatic. Asymptomatic infections are common in all malaria-endemic settings;^2^ although their prevalence is dependent on exposure to parasites, antimalarial immunity, use of preventive measures, and care-seeking behaviour.^3^ Asymptomatic infections with *Plasmodium falciparum* are particularly prevalent in school-aged children, who experience a higher force of infection and more persistent parasitaemia than younger children or adults.^4,5^ In persistent infections, parasite densities may fluctuate over time, and are often below the threshold of detection by microscopy or conventional malaria rapid diagnostic tests.^2,6^ These submicroscopic infections are disproportionately prevalent in areas of low endemicity and in settings that have experienced a recent decline in malaria burden.^2^

There is considerable debate about the characteristics of the human infectious reservoir and understanding the relative contributions of symptomatic and asymptomatic infections to onward transmission of malaria parasites to mosquitos is essential for guiding interventions to reduce and eliminate transmission.^7-9^ These contributions depend on infection density, infection duration, care-seeking behaviour, and importantly, gametocyte production. The likelihood that mosquitos become infected is positively associated with gametocyte density.^10^ However, due to variability in both the timing and level of *P. falciparum* gametocyte production, and the long gametocyte maturation process, gametocytes are absent during the early stages of a human infection. Thus, an association between asexual parasite and gametocyte density may be lacking in symptomatic infections and is weak in asymptomatic infections.^11-14^

To better understand the contributions of different populations to the human infectious reservoir for malaria, we performed a longitudinal study in Tororo, Uganda, previously a high transmission area that is currently under effective malaria control due to broad implementation of long-lasting insecticidal nets (LLINs) and indoor residual spraying of insecticides (IRS). We assessed the density, genetic composition, and kinetics of naturally acquired *P. falciparum* infections in an all-age cohort of Tororo residents. Gametocyte carriage and infectivity to mosquitos were quantified and compared between age groups and between participants with symptomatic and asymptomatic infections.

## METHODS

### tudy design and participants

The study was conducted in Nagongera sub-county (Tororo district) in eastern Uganda. Before 2013, malaria control in the district focused on malaria case management with artemether-lumefantrine, distribution of LLINs during antenatal visits, and promotion of intermittent preventive treatment during pregnancy. Universal LLIN distribution campaigns were conducted in November 2013 and May 2017. Sustained IRS has been maintained since December 2014, starting with rounds of bendiocarb every 6 months, and followed by annual rounds of pirimiphos-methyl (Actellic) since June 2016.^15^ Prior to IRS implementation, the annual *P. falciparum* entomological inoculation rate was 238 infectious bites per person per year (ib/p/yr) which was markedly reduced to 0.43 ib/p/yr five years later.^15^ In this context, we conducted a longitudinal study that is described in detail elsewhere. ^15^ Ethical approval for the study was received from the Uganda National Council of Science and Technology (HS119ES), Makerere University School of Medicine, the University of California, San Francisco and the London School of Hygiene & Tropical Medicine.

### Procedures

At baseline and at routine visits conducted every 4 weeks for 2 years, a standardized clinical evaluation was performed, and blood samples were collected. Cohort participants were encouraged to attend a dedicated study clinic open daily for all medical care. Study participants found to have a tympanic temperature > 38.0°C or history of fever in the previous 24 hours had a thick blood smear read urgently. If the thick blood smear was positive for malaria parasites by microscopy, samples were taken and malaria was treated according to national guidelines. Blood samples drawn at the onset of each malaria episode and at routine visits were assessed by quantitative PCR (qPCR) for *P. falciparum* parasite density, targeting the multi-copy conserved var gene acidic terminal sequence (varATS), with a lower limit of detection of 0.05 parasite/µL using 200 µL whole blood samples.^16^ For all qPCR positive samples, 100 µL of blood in RNA preservative (RNAprotect Qiagen) was used for automatic extraction of nucleic acids to quantify male (PfMGET mRNA transcripts) and female (CCp4 mRNA) gametocytes by quantitative reverse transcriptase PCR (qRT-PCR) with a lower limit of detection of 0.1 gametocytes/µL. ^17^ For the few (n=3) samples that were microscopy positive for gametocytes but qRT-PCR negative, microscopy gametocyte densities were used for analyses. Mosquito membrane feeding assays were performed on selected individuals who were diagnosed with symptomatic malaria, or at the time of routine visits if an individual was qPCR positive at their preceding visit, as described previously.^18^ Ten days post-feeding, mosquitos were dissected in 1% mercurochrome and examined for the presence of oocysts on the mosquito midgut wall by two microscopists; *P. falciparum* DNA (18S) was detected in infected guts by nested PCR. To assess parasite diversity, DNA from study participants and infected mosquito midguts was analyzed by amplicon deep sequencing of the apical membrane antigen 1 (AMA-1) gene, as described previously,^19^ over the entire period of follow-up. New infections were defined as the appearance of a new clone or group of clones at least 60 days after enrolment; to account for negative samples due to fluctuations in parasite density, we allowed 3 “skips” in detection, and classified an infection as cleared only if it was not identified in 4 sequential samples from routine visits^19^.Entomologic surveillance was performed throughout follow-up, with mosquito sampling every two weeks with CDC light-traps in all rooms where cohort participants slept, and detection of *P. falciparum* sporozoites in these mosquitos by ELISA, as described elsewhere.^15^

### Statistical analysis

Statistical analyses were performed in R, version 3.1.12; ^20^ full details on statistical analyses are presented in **supplemental methods**. Poisson regression with generalized estimating equations to account for repeated measures was used to estimate incidence of symptomatic malaria and of asymptomatic infections. Multiple generalized linear models were used to model measures of associations and present regression coefficients (β). Where appropriate, subject specific random intercepts were added to account for correlations between observations from the same individuals. For models with a dichotomous outcome or proportion response, we assumed a binomial distribution. A log link was used for proportion responses and a logit link used for dichotomous outcomes. All continuous densities (*d*) were log transformed as log_10_(*d* + 0.001) to allow densities that could possibly be zero to be retained in the model. When zero densities were not needed (e.g. for analyses including symptomatic malaria infections only), the transformation was log_10_(*d*) To model the relationship between total gametocyte density and proportion of infected mosquitos, we considered only *P. falciparum* positive episodes and used generalized linear regression models similar to the approach described by Bradley *et al*.^10^ The contribution of different populations to the infectious reservoir was estimated for i) symptomatic, asymptomatic microscopy-detected, and qPCR only-detected infections,^9^ and ii) ages <5 years, 5-15 years and ≥16 years. For the latter calculations, all episodes were included; for episodes in which no mosquito feeding assays were performed, the proportion of infected mosquitos was modeled using the relationship between gametocyte density and proportion of infected mosquitos (**supplemental methods**).

## RESULTS

Between October 4 and October 31, 2017, all 466 residents from 80 randomly selected households were enrolled into the study cohort; an additional 65 residents (total = 531) were enrolled when they joined participating households during the two-year follow-up period that ended on October 31, 2019. At baseline, parasites were detected by microscopy in 28 participants (5·3%) and by qPCR in an additional 64 participants (12·1%). Median age at enrolment was 8·7 years (IQR 3·7-26·4 years); 177 (33·3%) participants were <5 years, 193 (36·3%) 5-15 years and 161 (30·3%) ≥16 years of age.

Mosquito exposure was relatively low; on average two female *Anopheles* mosquitos were collected per sleeping room per night, with peaks following seasonal rains (**Figure 1A**). Of 15,780 female *Anopheles* mosquitos collected, 9 were positive for *P. falciparum* sporozoites (0·06%). During study follow up, in which 12,728 qPCR assays were conducted, a decline in the number of asymptomatic infections detected per month was observed. Overall, 38 episodes of symptomatic malaria were diagnosed over 955 person-years, resulting in an overall incidence of 0.040 episodes per person-year (0·048, 0·050 and 0·020 in participants aged <5 years, 5-15 years, and ≥16 years, respectively). During the same period, 110 new asymptomatic infections were detected, giving an asymptomatic infection incidence of 0.12 episodes per person year (0.07, 0.13 and 0.14 in participants aged < 5 years, 5-15 years, and >16 years, respectively). Mean parasite prevalence over the study period in participants aged <5 years, 5-15 years, and ≥16 years was 1·2%, 3·3%, and 0·8% by microscopy, and 4·0%, 14·0%, and 10·8% by qPCR, respectively (**Figure 1B&C)**. Geometric mean parasite density among symptomatic malaria infections was 10,825 parasites/µl (95% CI 5,419-22,763), markedly higher than parasite densities among asymptomatic infections (2.67 parasites/µl, 95% CI 2.22-3.17; **Figure 1D**). Among participants with asymptomatic infections, we observed lower parasite densities in the second year of follow-up, compared to the first, in individuals of all ages (mean difference in log densities between year 2 and year 1; <5 years: -0.51, 95% CI: -1.12 to 0.08, p=0.089; 5-15 years: - 1.03, 95% CI: -1.21 to - 0.83, p<0.001; and ≥16 years: - 0.35, 95% CI: -0.61 to - 0.09, p=0.008).

**Figure 1:**
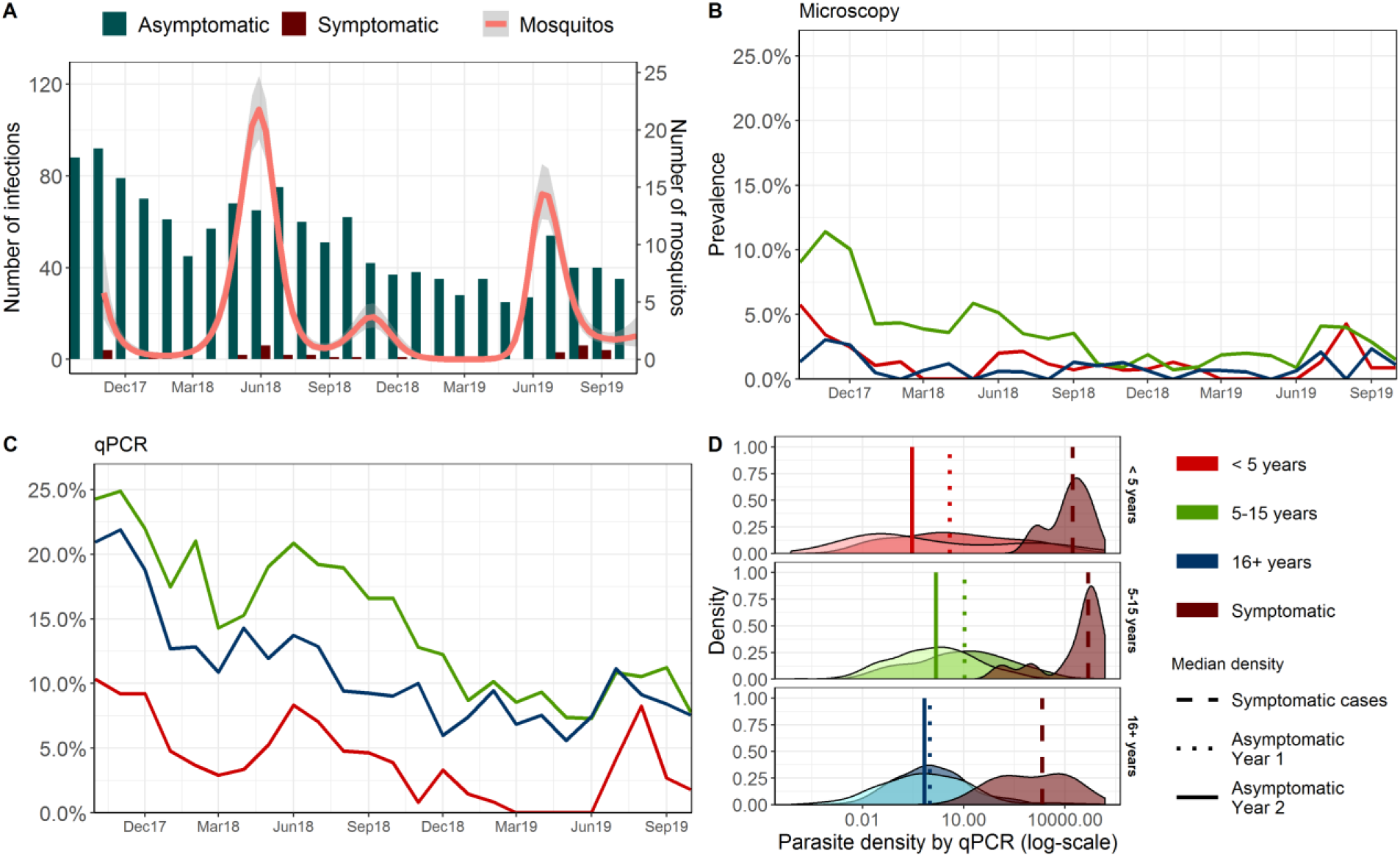
Symptomatic malaria episodes and asymptomatic *P. falciparum* infections. (A) Numbers of asymptomatic qPCR-detected infections, symptomatic malaria infections, and mean number of mosquitos caught per room per night. The red line for mosquitos represents a smoothed polynomial function (grey area is the 95% confidence interval). (B) Prevalence of *P. falciparum* by microscopy for individuals at indicated ages (legend bottom right). (C) Prevalence of *P. falciparum* by qPCR. (D) Parasite density distributions by qPCR for different age groups. Asymptomatic infections follow the colour scheme for age as defined in the legend, with vertical lines indicating median densities in the different years; the less numerous symptomatic infections are presented in maroon for the two years combined.

Overall, 538 mosquito feeding assays were performed with blood samples from 107 individuals diagnosed with either symptomatic malaria (24 experiments) or asymptomatic infections (514 experiments). Of the feeds performed on asymptomatic infections, 100 were done when individuals were parasite-negative, but with a positive qPCR result on the preceding visit (**Table 1**). Parasite densities among participants selected for mosquito feeding assays were similar to those measured in the entire cohort (**Supplemental Figure S1**). At least one infected mosquito was observed in 7·2% (39/538) of feeding experiments, with a mean of 1·2% (446/37,404) mosquitos infected per experiment (range 1·1-81·4%). Of experiments that resulted in infected mosquitos, 61·5% (24/39) were from asymptomatic microscopy-positive individuals; this population was responsible for 87·9% (392/446) of the infected mosquitos, with far fewer infected mosquitos arising from asymptomatic submicroscopic infections (11·2%; 50/446) or symptomatic malaria infections (0·9%; 4/446) (**Table 1**).

**Table 1.** Mosquito feeding results. A median of 69 (IQR 62-81) mosquitos were examined for infection status per feed. The proportion of all observations reflects the occurrence (of stated infection status and age group) among all observations (N=14,701).

| Age group | Infection status | Proportion of all observations in the cohort, % (n) | Proportion infectious feeds, % (n/N) | Proportion mosquitos infected, % (n/N) |
| --- | --- | --- | --- | --- |
| All | No parasites detected by qPCR | 77.5 (11,380) | 0.0 (0/100) | 0.0 (0/7,152) |
| <5 years | Sub-microscopic parasitaemia | 0.6 (94) | 0.0 (0/11) | 0.0 (0/618) |
| <5 years | Microscopically detected parasitaemia | 0.3 (37) | 75.0 (6/8) | 17.7 (86/486) |
| <5 years | Symptomatic malaria | 0.1 (12) | 0.0 (0/7) | 0.0 (0/484) |
| 5 – 15 years | Sub-microscopic parasitaemia | 3.9 (569) | 6.7 (14/206) | 0.4 (53/14,358) |
| 5 – 15 years | Microscopically detected parasitaemia | 1.2 (172) | 20.4 (17/83) | 5.2 (293/5,658) |
| 5 – 15 years | Symptomatic malaria | 0.1 (20) | 8.3 (1/12) | 0.5 (4/758) |
| >16 years | Sub-microscopic parasitaemia | 2.8 (405) | 0.0 (0/96) | 0.0 (0/6,879) |
| >16 years | Microscopically detected parasitaemia | 0.2 (32) | 10.0 (1/10) | 1.5 (10/667) |
| >16 years | Symptomatic malaria | 0.04 (6) | 0.0 (0/5) | 0.0 (0/344) |

Gametocyte prevalence by qRT-PCR was significantly higher among asymptomatic infections (63·8%; 884/1308) compared to symptomatic infections (28·9%; 11/38) (OR=3.2, 95% CI: 1.1-9.3; p=0.033). 46·0% (18/39) of infectious feeds and 61·2% (273/446) of infected mosquitos were from individuals with microscopically detectable gametocytes at the time of mosquito feeding. Amongst samples that were both parasite and gametocyte positive (n=895), gametocyte density was positively associated with concurrent total parasite density in asymptomatic infections (β=0.3, p<0.001), but not in symptomatic infections (β=-0.003, p=0.903) (**Figure 2A**). Gametocyte density was strongly predictive of the proportion of mosquitos that became infected when feeding on a blood meal (β=2.11, p<0.001; **Figure 2B**). Whilst there was no evidence for an interaction of age with the association between gametocyte density and proportion of mosquitos infected (β=0.088, p=0.113), gametocyte densities among gametocyte carriers differed between age groups (**Figure 2C**) (**Supplemental Figure S2**).

**Figure 2:**
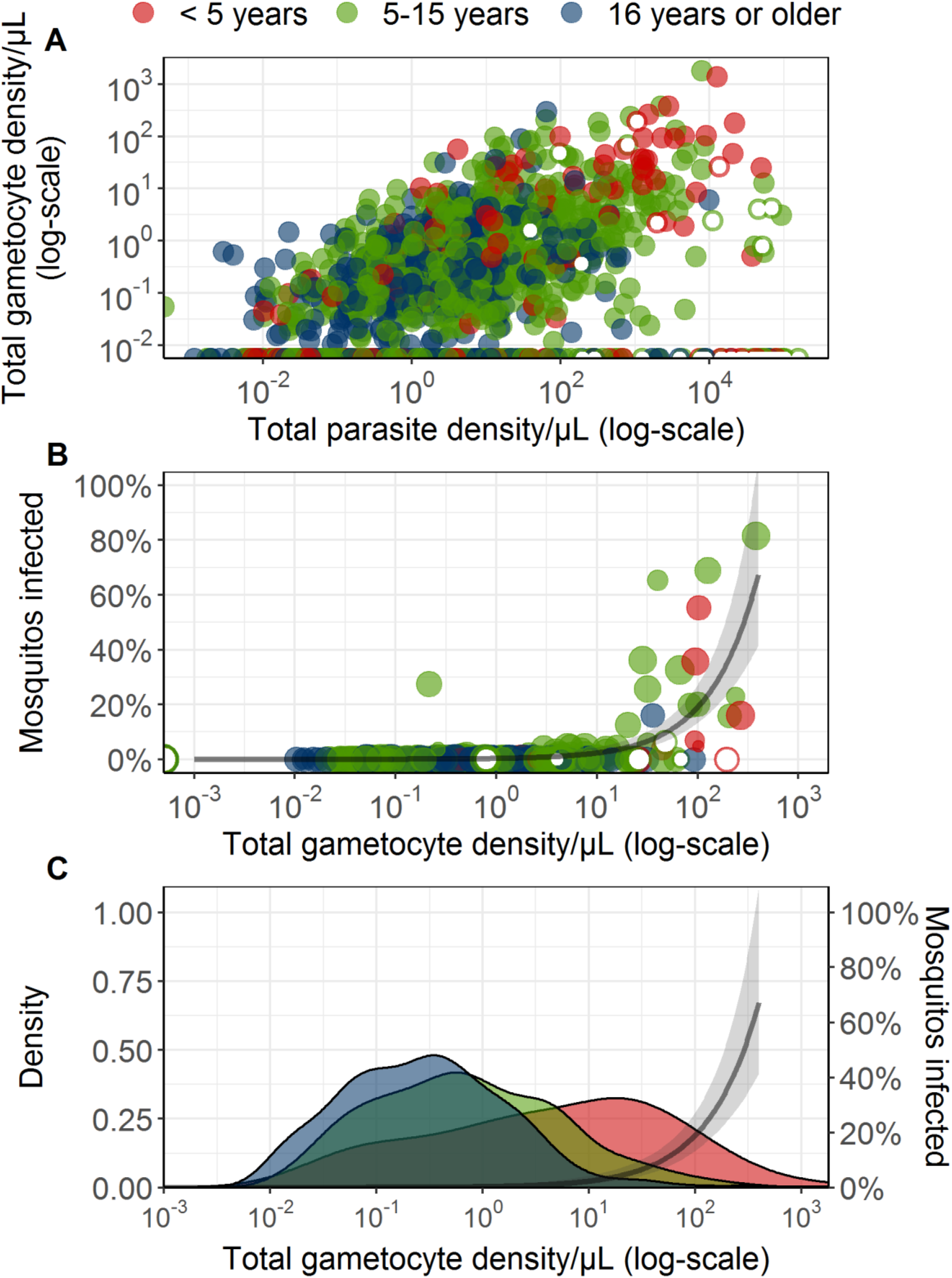
Gametocyte prevalence and density in relation to infectiousness to mosquitos. (A). Relationship between total parasite density and total gametocyte density. Each symbol is a parasite-positive episode; filled circles represent asymptomatic individuals and open circles symptomatic malaria infections. (B) Percentage of infected mosquitos in relation to gametocyte density. The size of the symbols reflects the number of mosquitos dissected; the colours are as in A. The line represents the best fitted association and the shaded area the 95% CI. (C) Gametocyte density by qRT-PCR among individuals who were gametocyte positive in different age groups (colours as above). Median gametocyte density among gametocyte carriers overall was 4.89 (IQR: 0.49 – 28.35) for children <5 years, 0.66 (IQR: 0.15 – 3.12) for children 5-15 years (p<0.001) and 0.28 (IQR: 0.07 – 0.89) for older individuals (p<0.001). The black line with grey shading is as in panel B.

Considering both transmissibility of infections and their relative frequency in our study cohort, asymptomatic microscopy-detected infections were estimated to be responsible for 83·8% of the infectious reservoir, with asymptomatic qPCR-detected infections and symptomatic infections being responsible for 15·6% and 0·6%, respectively (**Figure 3A**). We estimated the contribution of different age groups to the infectious reservoir based on mosquito feeding results and, when no feeding was done, parasite and gametocyte density measures to model infectiousness to mosquitos. We estimated that children aged 5-15 years were responsible for 56·8% of the infectious reservoir, followed by children aged < 5 years (27·5%) and participants age 16 years and older (15·7%) (**Figure 3B**).

**Figure 3:**
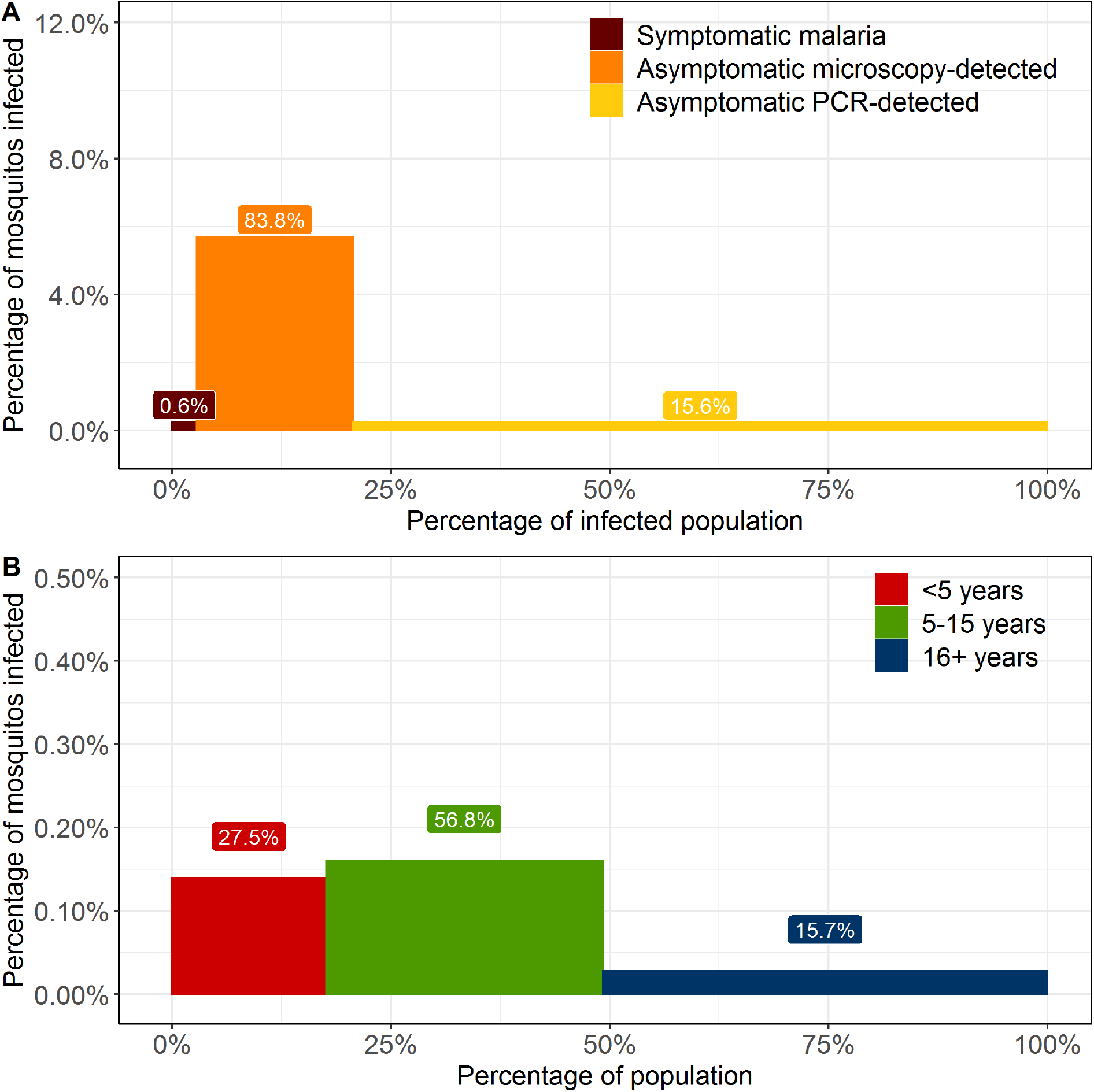
Contribution of different populations to the human infectious reservoir for malaria. (A) The contribution of different infection types to the infectious reservoir. Bar widths indicate the proportion of the infected population in each category; bar heights indicate the proportion of mosquitos that became infected when feeding on this population. Values above the bars are estimates of the proportional contribution of each of the three populations to the infectious reservoir. (B) The contribution of different age groups to the human infectious reservoir, displayed as described for panel A.

Overall, 75 individuals participated in multiple mosquito-feeding assays (range 2-19 assays), which allowed us to examine variations in transmission potential between individuals and over time during chronic infections. Whilst 35·9% (14/39) of infectious feeds were performed on samples from individuals with parasites below the level detected by microscopy, all infectious individuals were microscopy-positive for asexual parasites or gametocytes at least once during follow-up (**Figure 4A**). Remarkably, four children (0·8% of the cohort) were responsible for 62·6% (279/446) of all infected mosquitos. These highly infectious individuals were <5 years (n=2) or 5-15 years (n=2), some with clonally complex infections (up to 23 clones detected during follow-up); three of these children acquired infections during follow-up that were transmissible in the absence of symptoms. Twenty-eight parasitaemic individuals never infected mosquitos despite ≥5 mosquito feeds; 78·5% (22/28) of these individuals had infections documented by qPCR for more than 12 months (**Figure 4B**).

**Figure 4.**
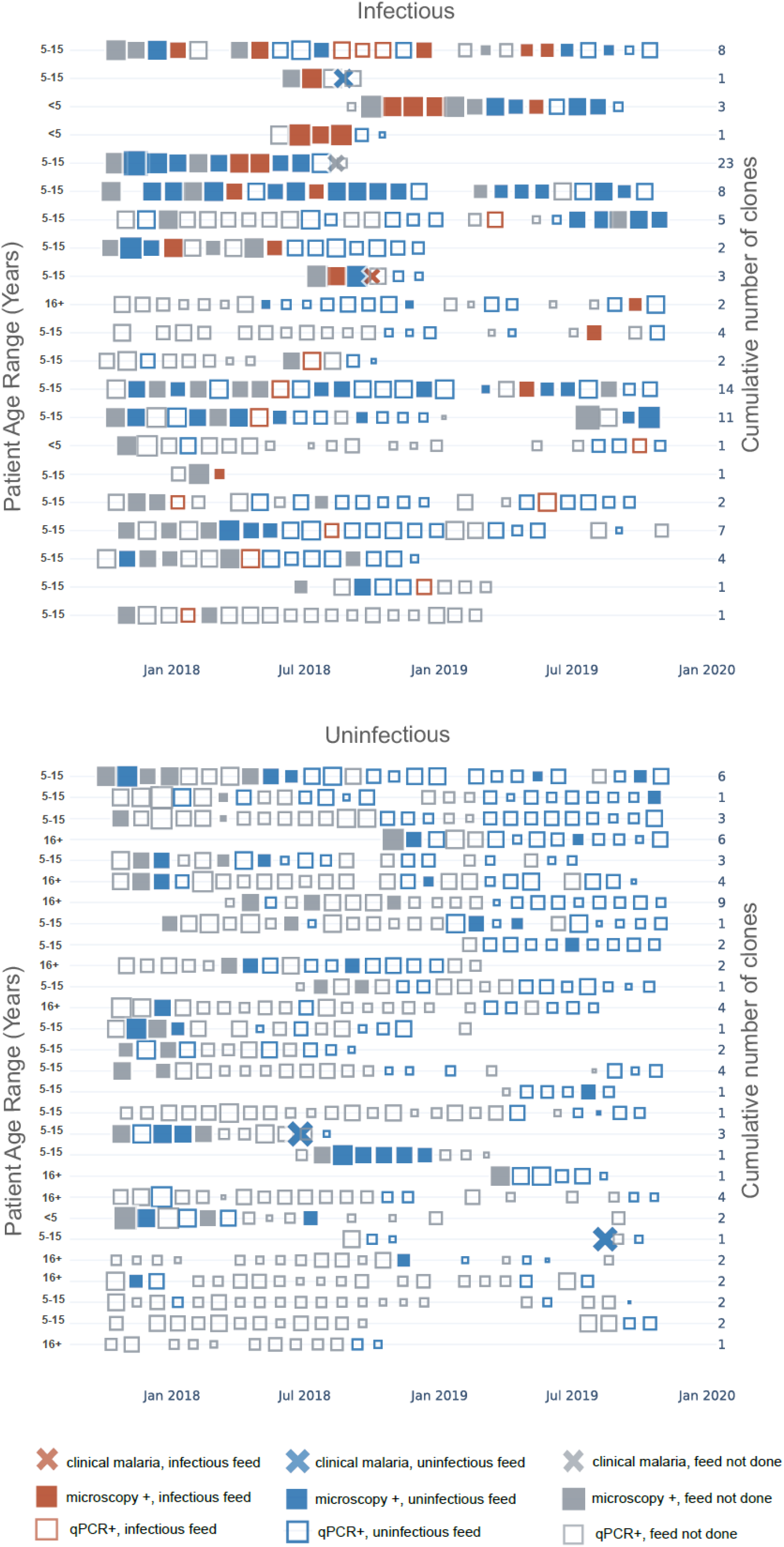
Longitudinal infectivity to mosquitos. Each row represents a cohort participant over the course of 24 months. Each square indicates a visit when parasites were detected, with the size of the square reflecting parasite density. The top panel (A) contains all individuals who were infectious on at least one occasion, ranked from top to bottom based on the total number of mosquitos they infected. The bottom panel (B) contains a selection of individuals (chosen from 86 individuals) who were never infectious but had repeated feeding assays.

*P. falciparum* DNA was detectable by nested-PCR in 92·8% (414/446) of infected mosquito midguts; amplicon deep sequencing was successful for 72·2% (322/446) of these samples, providing genotyping data from at least one mosquito for 33 of 39 (84·6%) successful feeds. Identification of genotypes over time, in relation to transmission to mosquitos, is presented for the four most infectious individuals in the cohort (**Figure 5**; graphs for other participants are provided in **Supplemental Figure S3**). Two genetically complex infections repeatedly transmitted multiple clones to mosquitos (**Figure 5A & 5C**); two single-clone infections resulted in short periods of high infectivity to mosquitos (**Figure 5B & 5D)**. Overall, 90·5% of parasite clones that were transmitted to mosquitos were detected in the blood of the participant on the day of mosquito feeding; 6·6% were only detected in the blood on prior visits, and 2·9% were never detected in the blood of that participant. Furthermore, there was a strong positive association between the abundance of a clone in peripheral blood and the abundance recovered in infected mosquitos (ß = 0.43, p <0.001) (**Supplemental Figure S4**).

**Figure 5.**
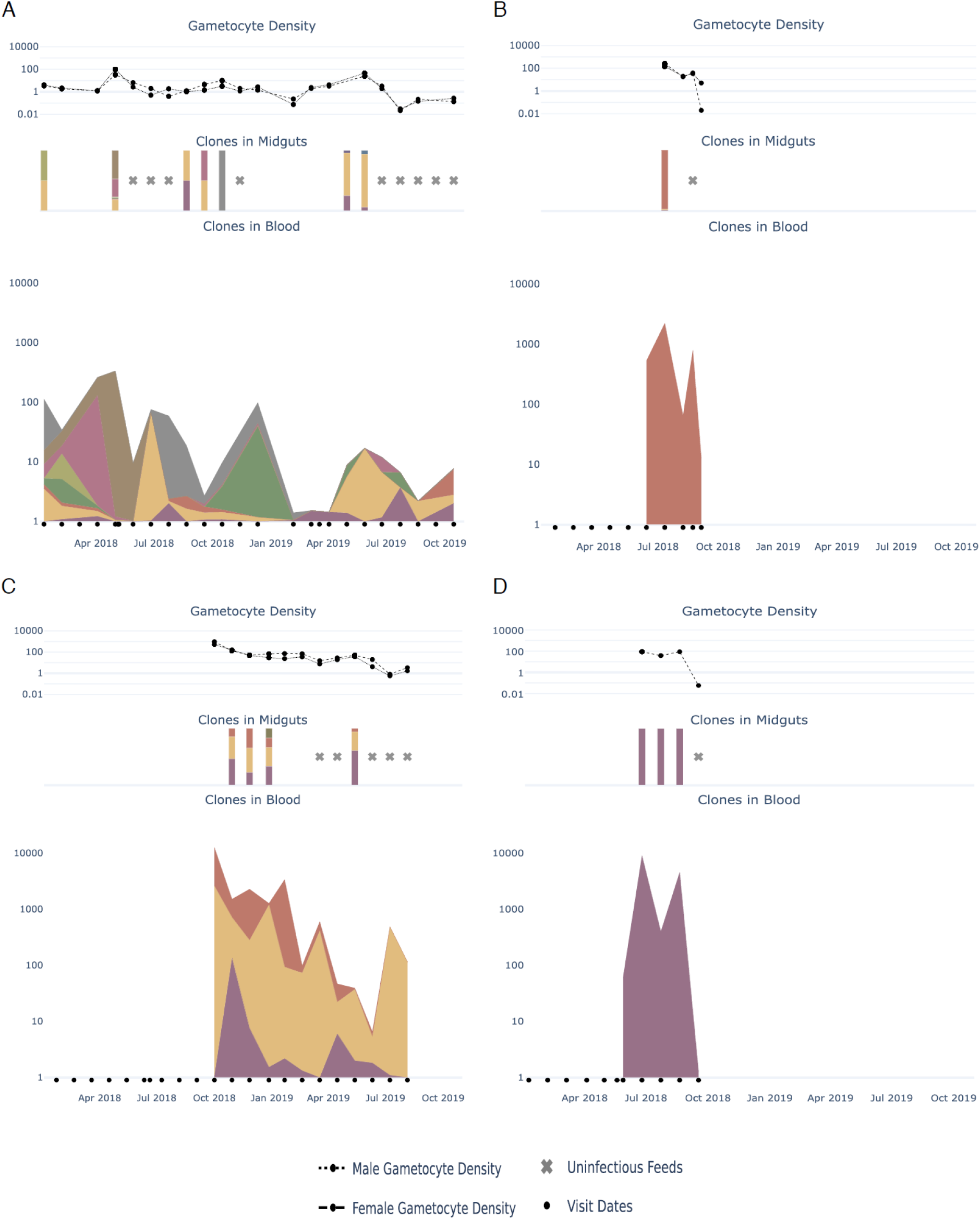
Gametocyte density and parasite clones recovered from blood and mosquito midguts in the 4 most infectious individuals. Male and female gametocyte densities and the number of clones detected in blood and infected mosquitos are shown for samples from the 4 individuals (A-D). Bars indicate infectious feeds. Total parasite density and clonal composition of blood samples are presented in the bottom panels. Each colour indicates the contribution of a unique *P. falciparum* clone

## DISCUSSION

To effectively eliminate malaria, the human infectious reservoir must be targeted. However, very few studies have quantified this reservoir, and those that did were largely cross-sectional in nature, ignoring the durations of asymptomatic infections and fluctuations in parasite densities over time.^2,6,21^ In this study, we determined *P. falciparum* infection status, gametocyte carriage and transmissibility over two years in an area of Uganda that previously had high malaria transmission, but is now under highly effective malaria control. Our longitudinal sampling allowed novel insights into natural transmission dynamics and their relevance for control. Asymptomatic microscopy-detectable infections, which were particularly prevalent in school-aged children, were the main source of mosquito infections. These asymptomatic infections commonly persisted for several months, during which they continued to produce infectious gametocytes. These findings highlight opportunities for demographically targeted malaria control interventions.

Asymptomatic parasite carriers comprised >99% of the human infectious reservoir for malaria in our setting. Within this asymptomatic population, parasite density was positively associated with gametocyte density,^22^ and those individuals with microscopically detectable infections were responsible for 83·8% of the infectious reservoir. Symptomatic malaria infections were uncommon (0·040 episodes per person year) and comprised only 0·6% of the human infectious reservoir. This very small contribution of symptomatic malaria infections to transmission is consistent with recent estimates for *P. falciparum* transmission in Ethiopia,^9^ but markedly different from findings in Cambodia and Thailand, where symptomatic malaria cases with high gametocyte densities were suggested to be more important than asymptomatic malaria infections for sustaining malaria transmission.^7,8^ These contrasting findings may, in part, be attributable to treatment-seeking behaviour. In our cohort, symptomatic malaria episodes were less likely, compared to asymptomatic infections, to be gametocytaemic upon presentation. This result suggests that most symptomatic malaria cases presented early, before the 9-12 day maturation of gametocytes was completed.^23,24^ In asymptomatic infections, parasite density, which is strongly associated with immunity of the infected individual, ^5,25,26^ is likely to be crucial in determining transmissibility. Parasite densities that were above the threshold of detection by microscopy were relatively common in cohort children, who frequently had concurrent gametocyte densities high enough to result in mosquito infection. Parasite densities generally decline with reduction in transmission intensity,^2,26^ and in some settings the majority of asymptomatic infections persist with ultra-low densities that are probably incompatible with transmission to mosquitos.^12^

The occurrence and kinetics of asymptomatic infections are related to immune responses that control parasite density and symptoms, and to genetic diversity of parasite populations.^2^ Whilst our study setting was historically characterized by intense transmission^15,27^, many of our cohort participants were born after IRS was implemented in the area and malaria exposure dropped to very low levels.^15^ Some of these young, presumably malaria-naive children acquired incident infections without experiencing symptoms, supporting earlier indications that children exposed to low transmission can acquire immunity very efficiently.^25^ However, several of these incident asymptomatic infections were highly important for transmission to mosquitos.

We observed marked differences in parasite carriage, parasite density and transmission potential between age groups. Parasite prevalence by microscopy and qPCR and parasite densities were highest among children aged 5-15 years.^4,5,28^ Older individuals had considerably lower parasite and gametocyte densities^4^ (**Supplemental Figure 2**). When combining the age distribution in the study district with parasitological and transmission data, individuals aged < 5, 5-15 and ≥ 16 years were estimated to comprise 27·5%, 56·8% and 15·7% of the human infectious reservoir, respectively. In all age groups, parasite densities showed a modest decline over time, with considerable fluctuations in densities during individual infections. In Vietnam, 7% of ultralow density *P. falciparum* and *P. vivax* infections escalated into microscopy positive infections.^6^ Longitudinal studies from Senegal and Ghana also showed marked fluctuations in the detectability of *P. falciparum* infections over time.^2^ We observed similar oscillations in parasite densities. A unique component of our study was longitudinal assessments of transmissibility to mosquitos of these oscillating infections (with up to 19 infectivity assessments per participant). We observed marked heterogeneity in transmission potential between and within individual infections, with four children (0.8% of our cohort) responsible for more than 60% of all infected mosquitos. These four children and all individuals who were infectious to mosquitos were microscopy-positive at least once during follow-up, suggesting that ‘test and treat’ approaches could be used for control. However, 15.6% of the human infectious reservoir was attributable to infections that were submicroscopic at the time of feeding. This suggests that, unless repeated frequently, ‘test and treat’ approaches will miss part of the human infectious reservoir.

Our study had several limitations. First, we used membrane feeding experiments that underestimate transmission potential compared to direct skin feeding.^29^ The lower sensitivity of membrane feeding was likely compensated by the large number of mosquitos we examined per feeding assay (median 69) that was much higher than natural mosquito exposure (average of 2 per indoor trapping night).^21^ Second, for logistical reasons we based our selection of participants for mosquito feeding on parasite carriage 4 weeks prior to feeds, and thus we did not measure infectiousness in the first weeks of incident asymptomatic infections. Third, health seeking behaviour among our cohort participants, who had exceptionally good access to care, may not have reflected care in other settings. Some of the symptomatic malaria infections that we detected early after infection would likely not have received such prompt treatment in other settings,and thus might have developed into chronic infections with transmissible gametocyte densities.

Our findings have several important implications for malaria control. We identified chronic asymptomatic infections with microscopically detectable parasitaemia as the most important drivers of transmission in our setting. These infections frequently occurred in school-aged children, supporting hypotheses that schoolchildren form an important and accessible reservoir that could be targeted by malaria control interventions, such as chemoprevention.^30^ Our finding that a small minority of individuals are disproportionally important for transmission poses challenges for community-wide interventions. Highly infectious individuals frequently carried parasites for several months at variable densities and without accompanying symptoms. Relying on passive detection of symptomatic infections or community mass screening with conventional diagnostics may thus have insufficient impact on the infectious reservoir for transmission in settings of declining transmission such as the one we studied here. Identifying these populations that likely sustain malaria transmission is key to progressing toward malaria elimination and to avoiding resurgence if regional malaria control deteriorates.

## Data Availability

Cohort data are available through a novel open-access clinical epidemiology database resource, ClinEpiDB. Data for the study conducted from October 2017 through October 2019 (referred to as PRISM2) can be found at https://clinepidb.org/ce/app/record/dataset/DS_51b40fe2e2

## Contributors

TB, CD, GD, SS, PJR, BG and MK conceived and designed the study. CA, JCR, JB, JO, AM, MC, PO, KL collected the data JR, CA, NT, JB, JCR and TB analysed the data. GD, IRB, JCR and LMK advised on the analysis. CA, TB, JB, PJR, CD, GD and SS drafted the first version of the manuscript. All authors saw draft of the manuscript, contributed to data interpretation and provided input on writing. All authors critically reviewed the manuscript.

## Acknowledgments

We thank the study team and the Infectious Diseases Research Collaboration for administrative and technical support. We are grateful to the study participants who participated in this study and their families.

