## Supplemental data and methods for "Persistent malaria transmission from asymptomatic children despite highly effective malaria control in eastern Uganda"

**Supplementary webappendix**

This webappendix formed part of the original submission and has been peer reviewed.
We post it as supplied by the authors.

Supplement to: Andolina C, Rek J, Briggs J et al. ***P. falciparum*** **gametocyte carriage and human infectivity to mosquitos: a longitudinal study in an area under effective malaria control in Uganda**

**Supplemental methods: Detailed statistical methods**

Statistical analyses were performed in RStudio.[1,2] For data manipulation and figures, we used the dplyr[3] and ggplot2[4] packages, and for analysis requiring repeated measures we used the geepack package[5]. For analysis requiring random effects, we used the lme4 and lmerTest packages.[6,7] Poisson regression with generalized estimating equations to account for repeated measures was used to estimate malaria incidence and incidence of asymptomatic infections. Any clones identified in the first 60 days of observation were considered to be baseline (persistent) infections. An incident asymptomatic infection was defined as any of the following after day 60 without associated symptoms: 1) any new clone (or group of clones) not seen in the patient prior, 2) a clone that had infected the patient before but had been cleared for 3 consecutive routine visits before re-infection, or 3) a positive qPCR result after three qPCR-negative visit.

Multiple generalized linear models were used to model the association between parasitological and infectivity outcomes and covariates(s). Where appropriate, subject specific random intercepts were added to account for correlations between observations from the same individuals. For models with a dichotomous or proportion response, we assumed a binomial distribution. Dichotomous responses were associated with a single Bernoulli trial. For proportion responses, we further specified the number of samples that contributed to this proportion being binomially distributed. A log link was used for proportion responses and a logit link used for dichotomous outcomes. For models with continuous responses, we assumed a gaussian distribution. All continuous densities $(d)$ were log transformed as $\log_{10}(d + 0.001)$ to allow densities that could possibly be zero to be retained in the model. When zero densities were not needed to be included in the model (e.g. for analyses including clinical malaria cases only), the transformation was $\log_{10}(d)$.

The general formula of the GLM used for the models was

$$g(y) = \beta' X + z_i + \epsilon_i \qquad (1)$$

where y is the response and g is the link function as described, X is the vector of covariates, $\beta$ is the vector of unknown regression coefficients (slopes) to be estimated, $z_i$ is the random intercept for the $i^{th}$ individual and $\epsilon_i$ denotes the error term. The random intercepts are assumed to be gaussian distribution with a mean of zero and some constant variance as estimated in the model, i.e. $\epsilon_i \sim N(0, \sigma_z^2)$. For continuous responses, $\epsilon_i \sim N(0, \sigma^2)$. In particular, to model the relationship between total gametocyte density and proportion infected, we considered only *P. falciparum* positive visits. The model fit, as generally described in (1), was a generalized linear regression model, where the proportion infected was assumed to come from a binomial distribution with n=number of dissections. A log-link function was used for g(.) and log transformed total gametocyte density ($\log_{10}(Tgam + 0.001)$) was used as X. The model for the estimated proportion ($\hat{y}$) infected was thus found to be

$$\hat{y} = \exp(-5.88 + 2.11 \times \log_{10}(Tgam + 0.001)) \quad (2)$$

This is similar to the model used by Bradley *et al.* [8]

The contribution of symptomatic, asymptomatic microscopy-detected, and PCR-detected infections to the infectious reservoir was estimated as the proportion of the infected population in each category weighted by the relative infectivity to mosquitoes of each category. The proportion of observations that was in each infection category was calculated based on all cohort data. The contribution of different age groups (<5 years, 5-15 years and ≥16 years) to the infectious reservoir was estimated as the estimated proportion infected mosquitos in each age category weighted by the proportion of the population in each age category. The proportion infected mosquitos was partially estimated. When mosquito feeding data was available, that estimate of proportion infected mosquitos was used. For *P. falciparum* positive visits without feeding assays performed, the proportion infected mosquitos was treated as missing data, and modelled using the relationship between total gametocyte density and proportion infected mosquitos from those with feeding assays (equation (2)). For all *P. falciparum* negative visits, the proportion infected mosquitos was specified as 0. We then found the average proportion infected mosquitos within each age category. To account for the variation from the model, and within the age groups, we re-ran the model 500 bootstrap samples and estimated the combined the standard errors within samples and between samples to compute 95% confidence intervals for the average proportion infected within each age category. The proportion of the population in each age category was based on UN population census data for Uganda. [9]

| | <5 | 5-15 years old | >16 |
|---|---|---|---|
| 2017 | 17,9% | 31,7% | 50,4% |
| 2018 | 17,6% | 31,6% | 50,8% |
| 2019 | 17,4% | 31,4% | 51,2% |
| mean | 17,6% | 31,6% | 50,8% |

**Supplemental methods: Determining copy numbers per gametocyte for CCp4 and PfMGET**

In order to determine the copy numbers per gametocytes for CCp4 and PfMGET, purified male and female gametocytes standards were generated and purified by FACS from PfDynGFP/PfP45mCherry reporter line parasites. Gametocytes numbers were quantified by microscopy using a Bürker-Türk counting chamber and verified by 18s qPCR. Synthetic RNA standards were ran against purified male/female gametocytes to determine copies per gametocyte for each marker (CCCp4 and PfMGET). Two different volumes of purified gametocytes were used for extraction and each sample was analyzed 3 times in duplicate in 3 independent PCR runs. Standards from 3 runs were combined and used to make an overall standard curve for each sample volume. Linear regression was performed on log transformed data to assess the relationship between the two measures (gametocytes/ml and copies/ml) and the y intercept indicated the number of copies per gametocyte(conversion factor). The conversion factor from copies to gametocytes/ml was 14 for PfMGET and 19 per CCp4. [10]

As low levels of CCp4 (10,000 rings equal to 1 female gametocyte) and PfMEGET (100,000 rings equal to 1 male gametocytes) are present in rings, background ring noise was calculated by dividing the absolute number of asexual parasites by 10.000 for female gametocytes and by 100.000 for male gametocytes. The background was subtracted from the actual number of female and male gametocytes detected by multiplex qRT-PCR. Samples with an estimated density <0.01 gametocytes per µl were considered negative.

**Supplemental Figure S1. Parasite density among the entire study cohort and the population selected for mosquito membrane feeding.**

A violin plot showing that the distribution of parasite densities among participants selected for mosquito feeding assays (red) were similar to the distribution of parasite densities among the entire cohort (green).

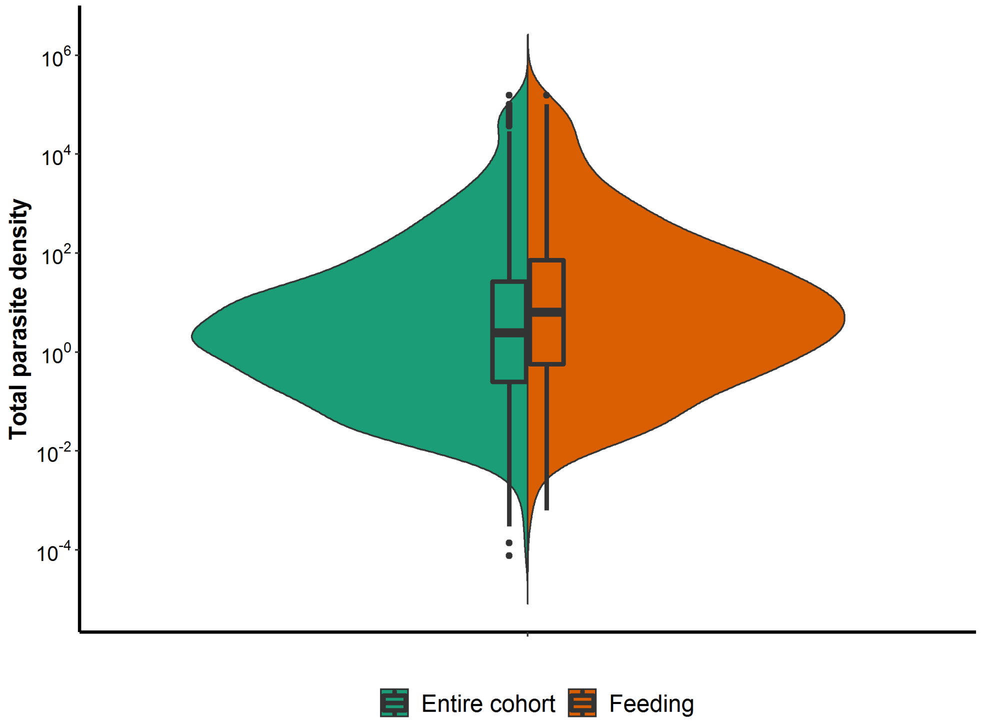

**Supplemental Figure S2. Parasite and gametocytes prevalence and density in relation to age.**

Parasite prevalence (A) and density (B) by qPCR are highest in children aged 5-15 years. Gametocyte prevalence (C) is lowest in children <5 years whilst among gametocyte positive individuals gametocyte density (D) is highest in this age group. Individuals aged ≥16 years often have gametocyte densities below the minimum density to allow mosquito infections. The black line with grey shaded area is the same as Figure 2B in the main file.

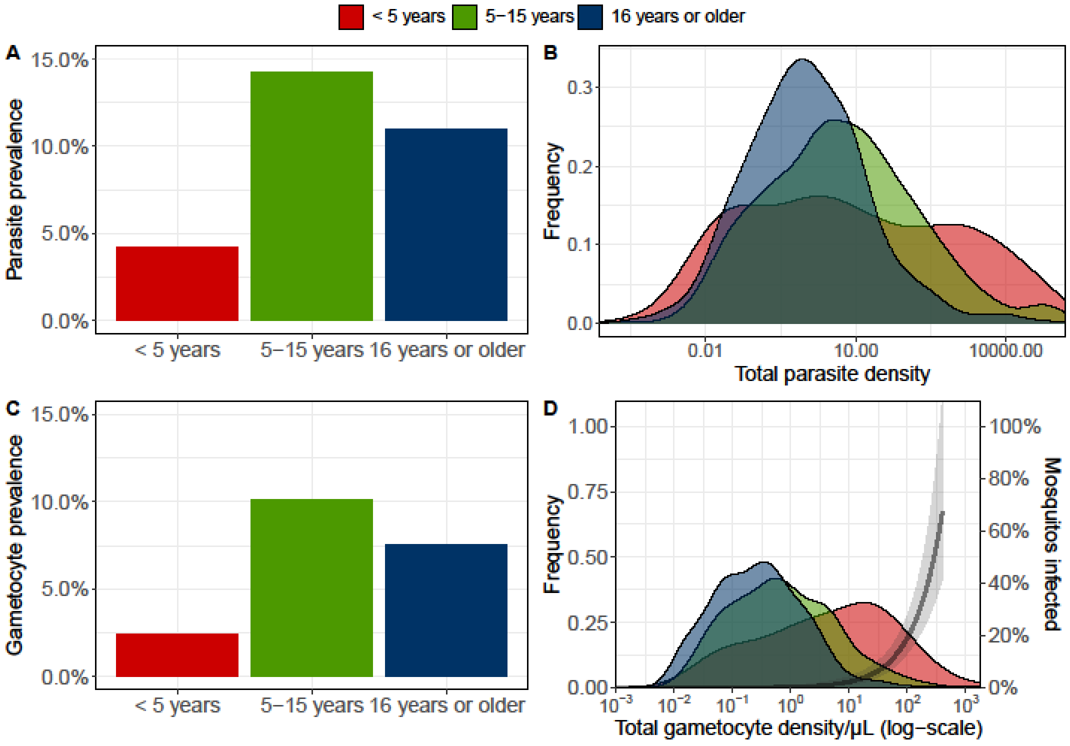


**Supplemental Figure S3. Gametocyte density and parasite clones recovered from blood and mosquito midguts in all infectious individuals.**

Male and female gametocyte densities and the number of clones detected in blood and infected mosquitos are shown. Bars indicate infectious feeds, with each color representing the proportion of each unique *P. falciparum* clone. Total parasite density and clonal composition of in blood samples are presented in the bottom panel with each color indicating the contribution of a unique *P. falciparum* clone.

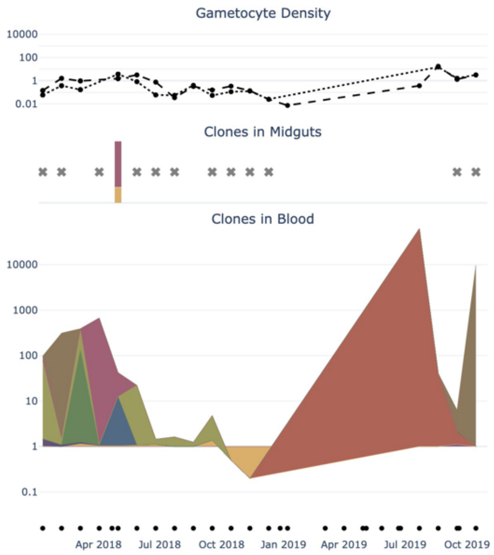
Gametocyte Density
10000
100
1
0.01
Clones in Midguts
Clones in Blood
10000
1000
100
10
1
0.1
Apr 2018
Jul 2018
Oct 2018
Jan 2019
Apr 2019
Jul 2019
Oct 2019

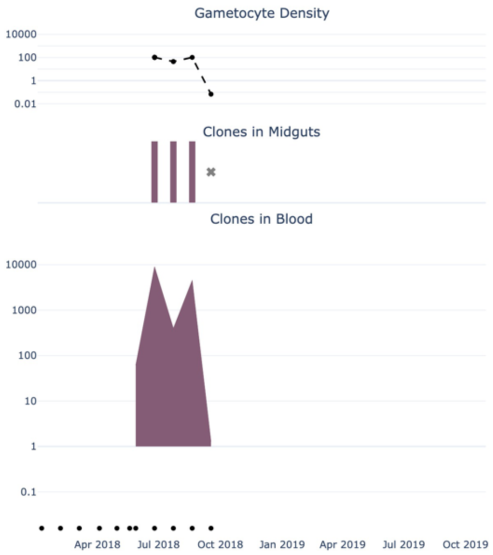
Gametocyte Density
10000
100
1
0.01
Clones in Midguts
Clones in Blood
10000
1000
100
10
1
0.1
Apr 2018
Jul 2018
Oct 2018
Jan 2019
Apr 2019
Jul 2019
Oct 2019

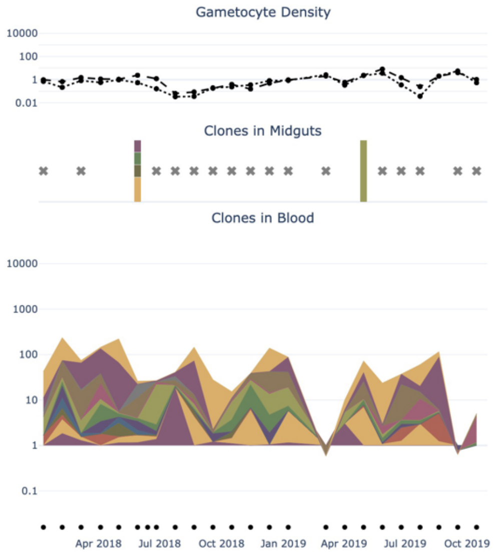
Gametocyte Density
10000
100
1
0.01
Clones in Midguts
Clones in Blood
10000
1000
100
10
1
0.1
Apr 2018
Jul 2018
Oct 2018
Jan 2019
Apr 2019
Jul 2019
Oct 2019

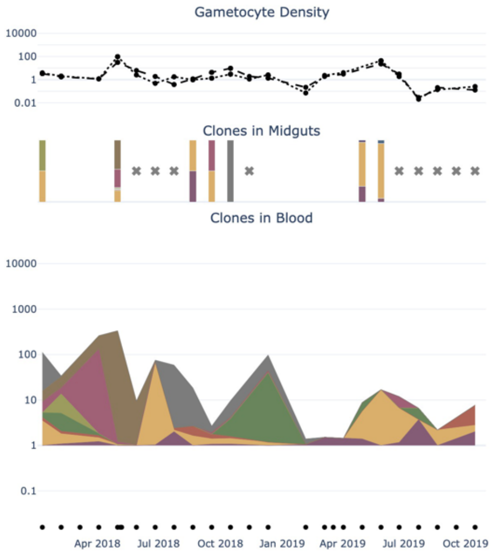
Gametocyte Density
10000
100
1
0.01
Clones in Midguts
Clones in Blood
10000
1000
100
10
1
0.1
Apr 2018
Jul 2018
Oct 2018
Jan 2019
Apr 2019
Jul 2019
Oct 2019

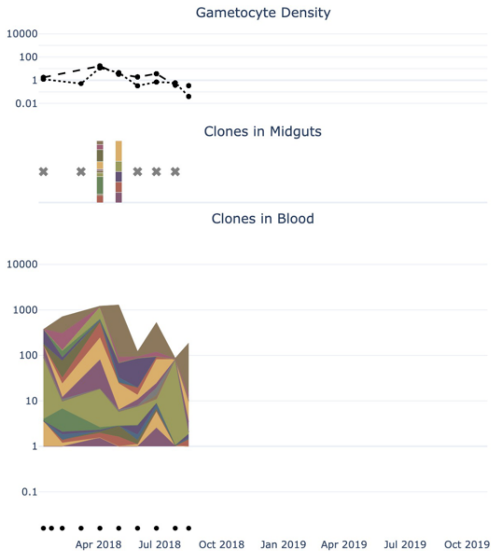
Gametocyte Density
10000
100
1
0.01
Clones in Midguts
Clones in Blood
10000
1000
100
10
1
0.1
Apr 2018
Jul 2018
Oct 2018
Jan 2019
Apr 2019
Jul 2019
Oct 2019

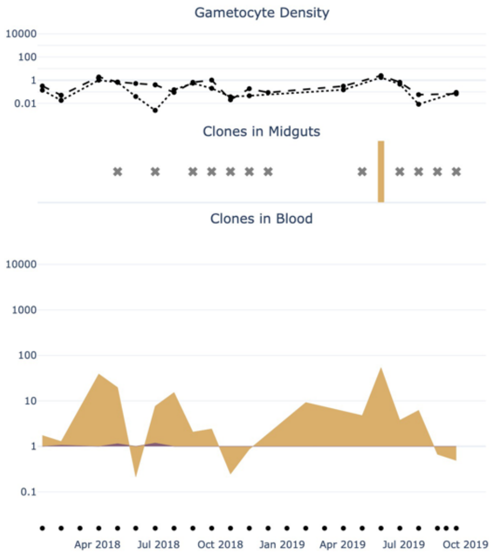
Gametocyte Density
10000
100
1
0.01
Clones in Midguts
Clones in Blood
10000
1000
100
10
1
0.1
Apr 2018
Jul 2018
Oct 2018
Jan 2019
Apr 2019
Jul 2019
Oct 2019

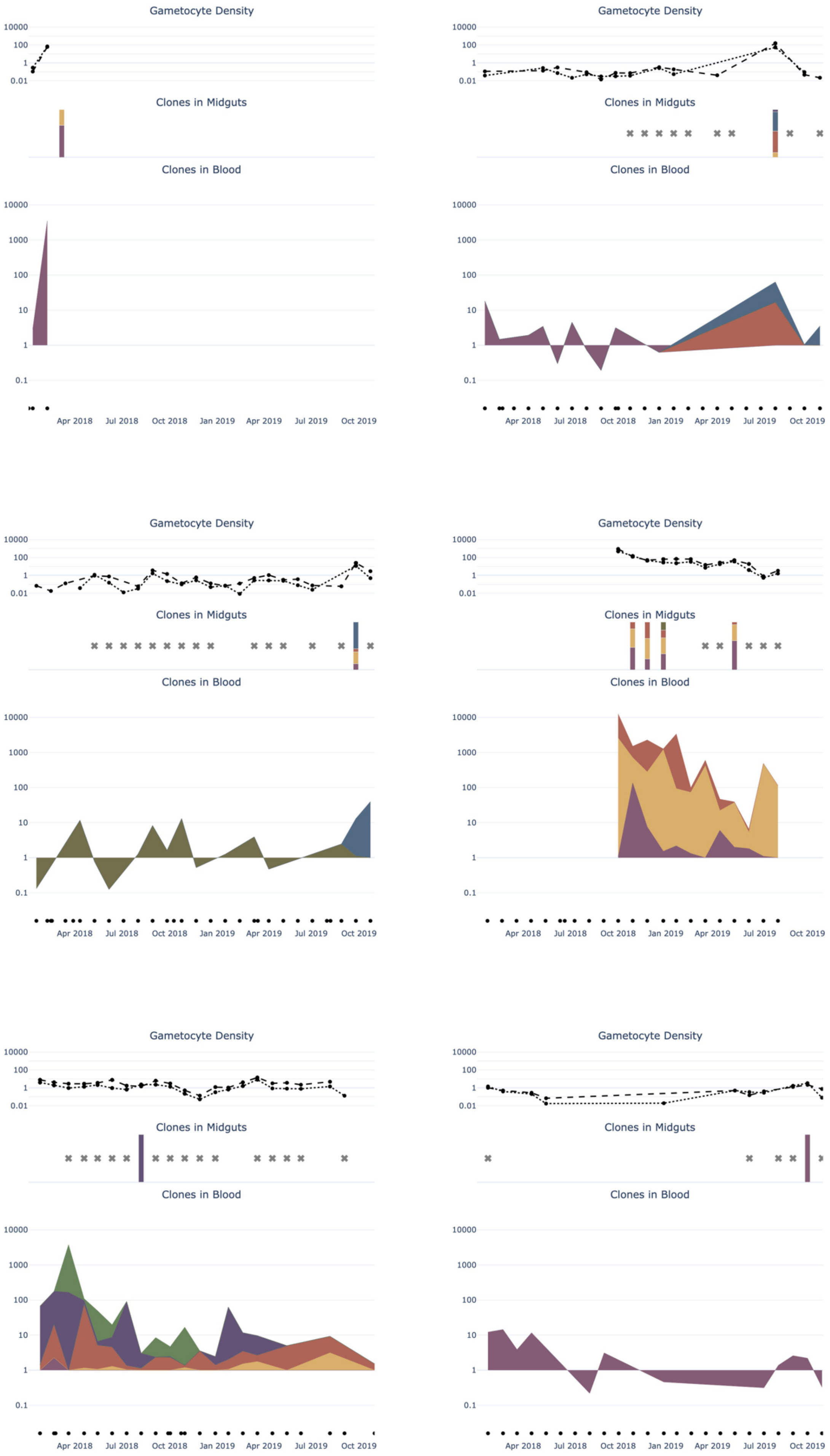
Gametocyte Density
10000
100
1
0.01
Clones in Midguts
Clones in Blood
10000
1000
100
10
1
0.1
Apr 2018
Jul 2018
Oct 2018
Jan 2019
Apr 2019
Jul 2019
Oct 2019

**Supplemental Figure S4. Abundance of a clone in peripheral blood versus the abundance among all clones recovered in infected mosquitos.** The blue line represents the regression of y ~ x with 95% confidence interval shaded in grey. Colours of dots indicate the clonal complexity in the blood sample.

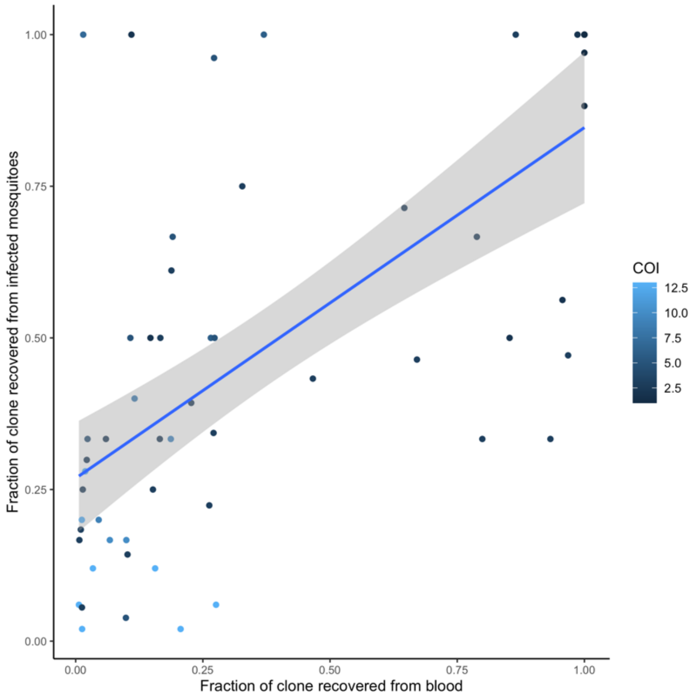
